# Development and evaluation of a behavioral intervention to reduce household plastic waste burning in rural Guatemala: Study protocol for Ecolectivos, a cluster-randomized community trial

**DOI:** 10.1101/2025.01.08.25320211

**Authors:** Lisa M. Thompson, Hina Raheel, Amy E. Lovvorn, Mayari Hengstermann-Artiga, Maria Renee Lopez, Alex Ramirez, Dana Boyd Barr, Melinda Higgins, John P. McCracken, Eri Saikawa, Margaret Handley

**Affiliations:** Nell Hodgson Woodruff School of Nursing, Emory University, Atlanta, GA, USA; Gangarosa Department of Environmental Health, Rollins School of Public Health, Emory University, Atlanta, GA, USA; Center for Health Studies, Universidad del Valle de Guatemala, Guatemala City, Guatemala; Department of Environmental Health Science, College of Public Health, University of Georgia, Athens, GA, USA; Department of Environmental Sciences, Emory University, Atlanta, GA, USA; Department of Epidemiology and Biostatistics, University of California San Francisco, San Francisco, CA, USA; PRISE Center (Partnerships for Research in Implementation Science for Equity), University of California San Francisco, San Francisco, CA, USA

**Keywords:** waste management, plastic, air pollution, implementation science, climate change

## Abstract

**Objective:** Household waste burning, especially of plastics, is a major, but unaddressed environmental and health hazard in countries that lack infrastructure to properly manage waste. This study will implement village-level community working groups that aim to reduce household plastic waste burning and improve health-related quality of life in women of reproductive age in rural Guatemala.

**Methods:** Using a type 1 hybrid-effectiveness-implementation study design, we will randomize 16 rural villages in Jalapa, Guatemala and randomly select 400 women of reproductive age (25 in each village) who report burning plastic trash as a primary form of waste disposal to participate. In eight intervention villages, we will conduct 12-week community working groups to implement alternatives to burning plastic that are achievable over the subsequent 9 months. We will use the Behavior Change Wheel and RE-AIM, two implementation science frameworks, and a mixed-methods approach, to refine, implement, and evaluate community-initiated interventions that address plastic waste. At baseline, 4 and 12 months, we will measure personal exposures to fine particulate matter and black carbon, and urinary biomarkers of exposure (e.g., bisphenols, phthalates, polycyclic aromatic hydrocarbons, and volatile organic compounds). We will use filter-based 1,3,5-Triphenylbenzene, a known tracer of plastic incineration, to quantify emissions estimates of air pollutants due to plastic burning. Based on plastic waste reductions in intervention villages, we will assess regional impacts of pollutant emissions reduction, using a 3D chemical transport model. This study was registered at ClinicalTrials.gov (NCT05130632) on November 23, 2021.

**Significance:** Our findings will be incorporated into community-driven public health actions to develop programs in other local contexts. This project has direct benefit not only to those residing in Guatemala, but potentially in other areas where open waste burning contributes to air pollutants both regionally and globally.

**Contributions to the literature:**

- The Ecolectivos project will engage communities to co-create sustainable alternatives to burning household plastic waste.
- Research will be refined during the formative phase, working groups will address behavior change during the main trial, and results will be disseminated using village-level fairs to introduce alternatives to burning plastic in control villages.
- This work will expand the evidence on barriers and enablers to implementing alternatives to burning plastic using the capability, opportunity, and motivation domains for key behaviors (guided by the Behavior Change Wheel framework) and focusing on reach and potential for scale-up (guided by RE-AIM framework).

## Background and Rationale

Household air pollution from solid fuel combustion (e.g. wood, crop waste, dung) is a major environmental risk factor in low-resource countries, accounting for an estimated 2.6 million deaths annually [1]. Increasingly, plastic is being burned either as kindling [2] or simply as a way to dispose of the insurmountable plastic waste in these settings. The exposure levels and health effects of pollution from plastic waste burned in household fires are understudied. While global efforts are introducing cleaner fuels and stoves as a replacement for solid fuels [3,4], these programs do not completely address plastic burned in household fires. This is challenging for the two billion people globally who lack municipal sanitation services [5]. In rural Guatemala, 71% of households burn waste, including plastic, as a primary means of disposal [6].

Open burning of plastic waste in household fires is a critical, yet understudied, public health and environmental hazard [7]. Combustion of plastic releases toxic smoke that contains carcinogenic [8] and endocrine-disrupting [9] compounds. While studies suggest that low levels of bisphenols and phthalates from plastics disrupt neurodevelopment [10], endocrine [11], and reproductive function [12], exposure to these compounds in women of reproductive age who burn their household plastic waste has not been reported in the literature. Furthermore, no intervention studies have been implemented in low-resource settings to reduce plastic waste burning. Finally, there are no estimates of ambient emissions from household-level plastic waste burning for air quality modeling in Central America.

Intervention strategies grounded in theoretical models that address the enablers and barriers to behavioral change can fill the implementation gap between what we know (evidence) and what we do (practice) [13]. One widely used approach to determining key areas for intervention targets and strategies to affect change, is the COM-B (‘Capability’, ‘Opportunity’, ‘Motivation’ and ‘Behavior’) model, part of the Behavior Change Wheel (BCW). Together, they provide a framework for assessing capabilities, opportunities and motivations, the conditions affecting key behaviors that must be understood contextually, before developing effective intervention strategies. Capabilities are demonstrated by applying necessary skills and knowledge. Opportunities are external social or environmental factors that inhibit or allow change. Motivation is guided by reflective or emotional processes that direct behavior. The COM-B Model is at the center of the BCW framework. Nine intervention functions address gaps in capabilities, opportunities and motivations that, when implemented correctly and consistently, will lead to sustained behavior changes [14]. The related Theoretical Domains Framework (TDF) is a synthesis of 84 theoretical constructs of behavior change organized into 14 domains [15] that maps directly onto the COM-B model and BCW and provides a more details characterization of the factors that affect the behavior(s) that is targeted by the intervention [16]. The BCW can be used to develop behavioral interventions but also to assess the delivery of implementation strategies for intervention components. Few studies have used the BCW for behavioral change related to community environmental health [17–20], but there is considerable recognition that theory-driven implementation science approaches, such as application of the COM-B/BCW frameworks are, necessary to identify promising intervention approaches that can be empirically tested [21].

In addition to applying approaches based on implementation science to environmental problems, methodological approaches that address the underlying drivers of pollution being disproportionately prevalent in places impacted by countries facing centuries of colonization[22], and subsequent structural and political marginalization of indigenous groups, as is the case in Guatemala, requires methodologies that prioritize community voices that have been historically silenced [23]. Community-level engagement is critically important, particularly in low-resourced settings, like Guatemala, where people have been intentionally excluded from institutions, laws or policies that protect them from environmental harms [24]. Addressing systemic problems of pollution builds necessarily on qualitative and/or mixed methods [25], environmental justice [26], health communication [27] and participatory research [28]. Feasible, implementable, and sustainable alternatives to the mounting plastic waste problem need to be prioritized in these communities.

## Study design and aims

No intervention studies have assessed exposure reductions of plastic combustion by-products, including bisphenols and phthalates, in women of reproductive age. Thus, we designed Ecolectivos, a village-level cluster randomized controlled trial (RCT) of a behavioral educational intervention (the community working group described below). Ecolectivos is a type 1 hybrid effectiveness-implementation study that measures the effectiveness of the community working group, while also evaluating how the intervention is implemented [29,30]. Our work is theoretically guided by two implementation science frameworks, (1) the COM-B model and the Theoretical Domains Framework (TDF) [16] and (2) the RE-AIM Framework [31]. The objective of Ecolectivos is to evaluate the adoption and sustainability of strategies to refuse, reduce, reuse/reintegrate, repair, and recycle plastic in anticipation of reductions in household-level plastic burning in intervention communities.

Our study aims are:

1. Implement a behavioral intervention (12-week community working group) and evaluate strategies chosen by the group that address plastic waste burning, targeting barriers and enablers identified within the capability, opportunity, and motivation domains, for key behaviors. The implementation outcomes assessed over 9 months will be reach, adoption, and maintenance/sustainability (implementation aim).
2. Evaluate the longitudinal effect of a behavioral intervention (community working groups) on urinary biomarkers of exposure to plastic waste burning (bisphenols, phthalates, polycyclic aromatic hydrocarbons and volatile organic compounds) and personal airborne fine particulate matter (PM_2.5_) and black carbon (BC) in 400 reproductive age women drawn from 8 intervention compared with women in 8 control villages. We <u>hypothesize</u> that biomarkers and exposures will decrease over time in women from intervention villages compared to women from control villages at 4-5 and 12-13 months (effectiveness aim).
3. Using 1,3,5-Triphenylbenzene (TPB) as a tracer of plastic burning and measurement of household plastic waste, apportion PM_2.5_ and quantify emissions estimates of air pollutants from plastic incineration and assess potential emissions reduction from the working group intervention on air quality with a chemical transport model.

## Materials and Methods

### Study setting

This 5-year study will be conducted in the rural mountainous region of Santa Maria Xalapán in Jalapa, Guatemala. The Jalapa Department has a rural population of 75,134 people in 133 villages. In this region, almost 50% self-identify as Xinca indigenous people; this proportion is higher in rural areas [6]. The remaining population self-identify as *Ladino*, or *mestizo*. In our earlier pilot work, 84% of women reported burning their household trash, including plastic [32]. We will seek formal permission from Indigenous councils and leaders of communities before recruiting participants into the study, as we have done in prior studies in the region [3].

### Formative research components (Year 1)

There were four main aspects of the formative research phase: 1) to form a community advisory board (CAB); 2) to conduct a baseline assessment survey; 3) to refine the working group procedures, including conducting a pilot working group in one community (Aim 1); and 4) to pilot exposure assessment methods for the Main trial (Aim 2).

### Eligibility criteria for formative phase

For the baseline survey and pilot working group, the inclusion criteria were: 1) age >18 years; 2) lives in study area; and 3) head of household and primary cook. Among the baseline assessment survey participants, selected 12 households that burned plastic trash inside and outside of their homes to participate in the air pollution monitoring pilot. For CABs and key-informant interviews, inclusion criteria were: 1) willing to attend quarterly CAB meetings and 2) identified as someone who works for a non-governmental or governmental organization on environmental protection or improvement, such as recycling, or a village leader.

We invited 12-15 people to form a Community Advisory Board (CAB), consisting of community stakeholders (e.g., councilmen from local villages, organizations working on waste management, recyclers, and governmental agencies (e.g., Ministry of Natural Resources and Ministry of Health)). The CAB was designed to provide input on study activities, evaluate working group intervention strategies, and discuss the potential to expand activities regionally. We extended formal invitations and met with individuals, but we were never able to form a CAB. Reasons for not joining the CAB included: individuals saw the CAB as a vehicle for personal gain, asking for large donations (e.g. a truck), those who worked for governmental agencies told us they were not authorized to work on projects that were outside their job scope, community leaders informed us that they hold their elected positions for a fixed term and could not commit to a 5-year membership, and indigenous leaders worked as a collective and did not want a single person to represent a communal voice.

The second formative phase activity was to conduct a baseline assessment of households in the Santa Maria Xalapán region. Using Google Earth imagery, geographical software, and the 2018 Guatemala Census to define a sampling frame and digitize building structures, we randomly selected 60 rooftops using satellite images from 37 selected villages with more than 200 households, oversampling to account for 30% misidentification of structures for a total of 2,220 potential households. Trained local fieldworkers used hand-held GPS receivers to identify 1,630 households. They verbally consented and administered a 30- minute survey to the primary cook about: (a) household size/composition; (b) stove type(s) and location(s); (c) location and frequency of burning waste, including plastic; (d) other types of waste that are burned in and outside of home; (e) recycling practices; (f) opportunities to dispose of waste besides burning; (g) motivation to improve the environment; (h) interest in participating in future community working groups; and (i) contact information. We identified participants for the Main Trial from these households.

Third, we piloted and refined the community working groups, including potential intervention strategies, like soap-making, recycling, and composting that can be implemented at the end of the working groups. We implemented the working group curriculum and tested all procedures in one village to standardize the scripts and materials. To refine the curriculum, we observed participants who burned plastic trash in their households, conducted open-ended surveys and key informant interviews with community stakeholders who recycle, dispose of, or repurpose plastic trash.

Finally, to test the methods for the Main trial, we conducted personal air pollution monitoring of plastic trash burning in 12 houses identified in the baseline assessment who reported burning trash in household fires. We monitored 24-hour personal and kitchen concentrations of personal airborne 24-hour fine particulate matter (PM_2.5_) and black carbon (BC) to estimate personal exposures and measured open waste burning over 1-2 hours. To assess two different tracers of plastic combustion, we measured filter-based 1, 3, 5-triphenylbenzene on 37 mm quartz filters and antimony (Sb) on 37 mm polytetrafluoroethylene (PTFE) filters[33–35].

## Main trial research components (Years 2-5)

### Eligibility criteria for main trial

To be eligible to participate in the Main trial, inclusion criteria are: (a) women of reproductive age (15 to < 44 years, verified by official document); (b) willingness to attend weekly meetings of the 12-week working groups if randomized to the intervention group; (c) willingness to participate in exposure/biomonitoring study; (d) reports using biomass as primary fuel for cooking; (e) reports daily participation in household cooking (does not need to be the primary cook) and (f) reports that plastic is burned in household fires (in cooking stove and outdoors) at least once a week. Exclusion criteria for women in the exposure/biomonitoring group are: (a) inability to consent; (b) individuals with impaired decision-making capacity; (c) currently pregnant, and: (d) women who report using tobacco products.

To be eligible to be an environmental health promotor, in addition to being in the Main Trial, participants must be over the age of 18 years and know how to read and write. For invited participants who attend the community-level working groups, there are no inclusion or exclusion criteria, but questionnaires administered to attendees will be limited to those over the age of 15 years.

## Intervention description

The community working group is a behavioral intervention with the goal of finding alternatives that lead to reduction or elimination of household plastic waste burning. The COM-B model is used in the intervention materials (e.g. by participants using take-home worksheets, by researchers using evaluation materials) to identify and subsequently address the capabilities, opportunities, and motivations to stop burning plastic trash that form the behavioral intervention. Participants over the age of 15 from the intervention villages will be invited to participate in 12-week working group sessions in a local village meeting place (**Table 1**). We will seek permission from village leaders and ask for their support in promoting working group activities, since from our experience they are decision-makers about village-level change. Two to four local Guatemalan fieldworkers will implement the 12-week working group which is grounded in the ecological and health implications of exposure to air pollution from burning plastic waste.

**Table 1:** Working Group Curriculum Themes and Components.

| Week | Theme | Components |
| --- | --- | --- |
| 1 | Identification of main problems of solid waste management, including sources of contamination in communities | Uses of plastics, plastic in waterways, plastic burning, introduction to recycling<br>*Homework:* Turn in pre-test; tally the amount of plastic in the household |
| 2 | Personal, family, and community practices of plastic waste management and effects on the ecosystem | Discussion of sources of contamination in the ecosystem and ways to reduce plastic use<br>*Homework:* Plastic production and marine life |
| 3 | Plastics in waterways and oceans, including discussion of microplastics; | *Group activity:* Make a stool out of plastic bottles and fabric |
|  | introduce the 4 R's (refuse, reduce, recycle, repurpose) | *Homework:* Time needed to degrade of plastic and other materials in the environment |
| 4 | Contaminants in burning plastic and health implications of exposure; discuss the difference between junk food in plastic bags and food like fruit, and vegetables, typically not wrapped in plastic | Discussion of alternatives to burning plastic; video on junk food versus healthy food<br>*Group activity:* Make worm composting containers<br>*Homework:* Find substitutions for plastic items used in the home |
| 5 | Sustainable alternatives to plastic and reducing plastic litter in the community | *Group activity:* Make organic soap (free of plastic)<br>*Homework:* Specify type and amount of plastic used during the week |
| 6 | Recycling plastic materials; discuss materials that can and cannot be recycled | *Group activity:* Sort recyclable plastic materials; Local recycling company comes to buy materials<br>*Homework:* Find items in home that are recyclable |
| 7 | Environmental justice; community empowerment | *Homework:* identify community problems that need to be addressed |
| 8 | Collective action; importance of community groups; describe plastic ban in Guatemala | *Group activity:* Community clean-up<br>*Homework:* Administer post-test; reflect on activities that participant wants to work on |
| 9-12 | Group selected 1-2 community activities, or “interventions”; this includes weekly meetings with Ecolectivos fieldworkers and community stakeholders who can support these activities. Project resources will be dedicated to implementing the activities, addressing bottlenecks to success. |  |

The Spanish-language manual (and English translation) used to develop the modular materials (PowerPoints, handouts and other didactic materials) is available at https://ecolectivosguatemala.org/material-educativo/. Eight core modules (weeks 1-8), the “essential ingredients”, and four additional modules (weeks 9-12) will be implemented weekly. During weeks 9-12, participants will identify intervention activities to reduce plastic waste burning. Participants will brainstorm on individual and community strategies that are important to them and will prioritize one or more strategies that can realistically be implemented over the next 9 months, under the guidance of Ecolectivos fieldworkers and designated promotors. These strategies may consist of, for example: 1) conducting community-level clean-up campaigns; 2) starting organic compost free of plastic waste; 3) training on community recycling, focusing on plastics; 3) making organic soaps, which can be used for personal grooming and washing clothes, eliminating plastic packaging; and 4) creating materials out of plastic, like soft drink bottle planters. Ecolectivos fieldworkers and promotors will work with community members to overcome bottlenecks to implementation during this period. We aim to maintain adherence (#11c) to core modules through strict protocols, while permitting flexibility of the selected activities, to allow integration of prioritized activities into community practices over the 9-month period [36]. Strategies to improve adherence to the behavioral intervention will include phone calls and home visits by our field team and promotors to encourage working group attendance, use of an attendance card stamped at each visit to monitor attendance, and in-class and take-home assessments about plastic waste, including waste burning in household fires.

## Outcome measures

Our **primary outcomes** are changes from baseline in personal airborne 24-hour fine particulate matter (PM_2.5_) and black carbon (BC) and changes in urinary concentrations of bisphenols (BPAs), phthalates, polycyclic aromatic hydrocarbons (PAHs) and volatile organic compounds (VOCs) (effectiveness, Aim 2).

Our **secondary outcomes** are implementation aims (Aim 1) and include reach, effectiveness, adoption, and maintenance/sustainability of the intervention. We will assess the effectiveness of the intervention implementation by asking women participating in the exposure monitoring at baseline and 12 months “do you burn plastic waste in your home?”. The RE-AIM evaluation framework is elaborated upon in **Table 2**.

**Table 2:** Exemplar of COM-B and RE-AIM frameworks to evaluate individual and community-level behavior change.

| Examples of COM-B Inquiry | Effectiveness Outcomes | Reach, Adoption and Maintenance Outcomes |
| --- | --- | --- |
| **Psychological/Physical Capability** |  |  |
| Identify plastic items in their home. (TDF domain=*Knowledge*) | N & % participants separating plastic and other recyclables correctly | % households report recycling by material type (*adoption*); variation in recycling by village (*reach*; *maintenance*) |
| Have you burned household trash? Plastic? (TDF domain=*Knowledge*) | N & % participants stating benefits from not burning plastic waste | % households de-adopting burning of plastics (*adoption*) |
| If you started an activity (like recycling or community clean-up), what would you need to be successful? (TDF domains =*Knowledge*; *memory*, *attention & decision processes*; *environmental context & resources*) | N & % participants still participating in the activity | % still participating in the activity (*adoption*); variation in participation by village (*reach*; *maintenance*) |
| If your household stopped burning trash/plastic, what would happen? (TDF domain=*Outcome Expectancy*) | N & % participants no longer burning plastic in indoor fires/in outdoor fires | % of villages no longer burning plastic (*adoption*); variation in burning by village (*reach*; *maintenance*) |
| **Reflective/Automatic Motivation** |  |  |
| Have you talked with your family members about not burning plastic? What have they said? (TDF domain=*Social Influences*) | N & % participants reporting that participation in educational sessions has improved their status within the home | Variation in perceptions by village (*reach*; *maintenance*) |
| Have you seen less plastic trash? Less plastic trash burning in your community? (TDF domain=*Optimism*; *Beliefs about consequences*; *Emotion*) | N & % participants who perceive less trash present/burning as part of community goals | Variation of villages attributing stopping burning trash for community goals by village (*reach*; *maintenance*) |
| Does burning plastic harm health? What happens if we don't take action to reduce plastic? (TDF domain= *Beliefs about consequences*) | N & % participants reporting health consequences from exposure to burning plastic | Variation of villages attributing stopping burning trash to health outcomes by village (*reach*; *maintenance*) |
| Is recycling a priority? Is not burning trash/plastic a priority? (TDF domain=*Social Influences*; *Goals*) | N & % participants and community members active during community clean-ups | Variation in villages participating in a municipal waste program and |
|  |  | reduced trash burning (*reach; maintenance*) |
| **Physical/Social Opportunity** |  |  |
| What community resources are needed to have a village “free of plastic”? (TDF domain=*Environmental context and resources*) | N & % accessing materials needed to create a create an improved environment in their village | Variation in villages accessing materials (e.g. recycling bins, clean-up resources, etc.) (*adoption, reach*) |
| Do people in your community care about trash or trash/plastic burning? (TDF domain=*Social Influences; Goals*) | N & % stating others have reduced trash/plastic burning | Variation in reporting by village (*adoption*), and by age, gender, etc. (*reach*) |

**Other outcomes** include quantification of emissions estimates of air pollutants from plastic incineration using filter-based 1,3,5-Triphenylbenzene (TPB) as tracers of plastic burning and collecting household plastic waste (Aim 3). Because engagement in a community working group intervention may affect women’s sense of well-being, we are measuring changes in Health-related Quality of Life Score, Household Decision Making Score, New General Self-Efficacy Scale, Short Social Capital Assessment Tool Score, and Community Mobilization Scale Score.

Outcomes will be measured at baseline (before randomization), at 4-5 months and 12-13 months (follow-up) in women who participate in the exposure/biomonitoring study. The follow-up period for both intervention and control participants included in this study will be about 13 months. Each matched community pair will be followed during the same period to control for seasonal and secular trends in local migration to harvest coffee, agricultural waste burning, and plastic waste burning. See **Figure 1** for the trial schedule.

**Figure 1.**
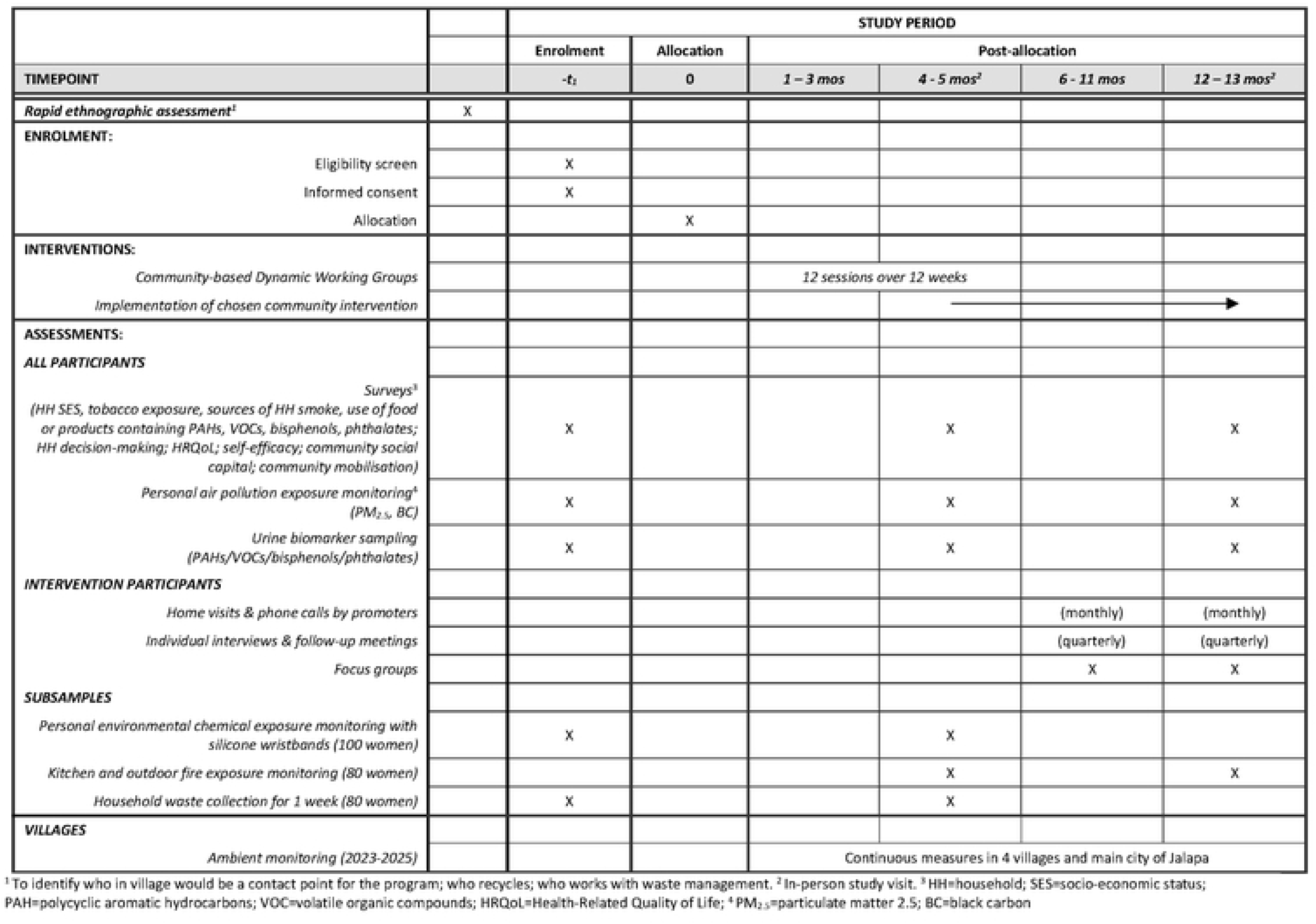
Ecolectivos trial schedule of enrolment, interventions, and assessments

### Sample size

Of the 400 women recruited for Aim 2, we estimate 10% will drop out by 12 months. We will ask women at baseline and 12 months “do you burn plastic waste in your home?” We assume that in control households there will be a 5% decrease in burning plastic waste. Using a cluster randomized design, and an estimated intraclass correlation coefficient of 0.2 for participants in the same village, we would be able to detect a 30% reduction in plastic burning in households in the intervention villages with 80% power and an alpha level of 0.05.

### Recruitment for Main trial

We will recruit 400 women of reproductive age from 16 villages from the baseline assessment conducted in the formative phase. Trained local Guatemalan fieldworkers will re-visit the women who expressed interest in participating in the Main Trial during the formative research survey. If women are not home after making three visit attempts (e.g., due to permanent move or temporary migration), or do not wish consent to participate in the Main Trial, the fieldworkers will choose the closest house to the left (when facing the front of the house), and so on, until an eligible participant is identified. This will be done until we enroll 25 women from each village who want to participate in the exposure monitoring study. Recruitment will be staggered over a 2-3 year period as each pair of villages enters into the study. Anticipated recruitment will be completed by January 2025.

### Additional recruitment

In addition to inviting women to participate in the Main trial, we will ask other village members to attend the working groups. These women may be neighbors or friends of the recruited woman, members of city councils, teachers, or church members. The goal is to have 50 people participating regularly in the community working groups. All participants in the working groups will provide written informed consent.

### Assignment of interventions

To conceal study arm allocation, the study manager at Emory University will prepare eight sets of numbered, sealed opaque envelopes (and one sample set) containing intervention/control assignments. The outside of each envelope will contain a label for recording the date of randomization; village name; group assignment (control or intervention); and the name of the person conducting the randomization. Envelope flaps will be sealed with a tamper-safe sticker. Each envelope will contain two identical folded pieces of paper, labeled either ‘intervention’ or ‘control’. Envelopes will be stored in sequence number order in a secure location at the study office. On the day of randomization, a Guatemalan fieldworker will check out the next sequential set of envelopes, and a “sample” set of envelopes, documenting this on a Randomization Envelope Log.

### Village-level randomization

Based on the baseline assessment survey, we will select eight pairs of non-contiguous communities matched on village population size, proximity to a main road, and distance from the city of Jalapa. Using a randomized cluster trial design, we will randomly select one village within each matched pair to receive the community working group intervention to implement strategies to reduce burning of plastic waste. Randomization will occur at a community meeting with representative leaders from both villages. We will review the study goals, process of randomization, and activities that will occur in both intervention and control villages during the study period. A fieldworker will show the contents of two unsealed sample envelopes, one containing a control assignment, the other an intervention assignment. They will then lay the two actual randomization envelopes on a table and each representative will choose an envelope and open it to reveal the assigned study group. One sheet of paper will be given to the village representative; the other will be placed back in the envelope. The fieldworker will then fill out all items on both randomization envelope labels. The envelopes will be returned to the study office, documented on the Randomization Envelope Log, and filed in a secure, designated location. Given the nature of the intervention, the participants and the fieldworkers will not be masked. Data analysts will be masked.

## METHODS: DATA COLLECTION, MANAGEMENT, AND ANALYSIS

### Data collection

All measures will be collected from 400 women who participate in the Main trial at baseline (before randomization), and at 4-6 months and 12-13 months from baseline during household visits and are further described in **Figure 2**. Visits will be conducted in the participant’s home after they have provided written informed consent. Data collection will be completed by May 2026.

**Figure 2:**
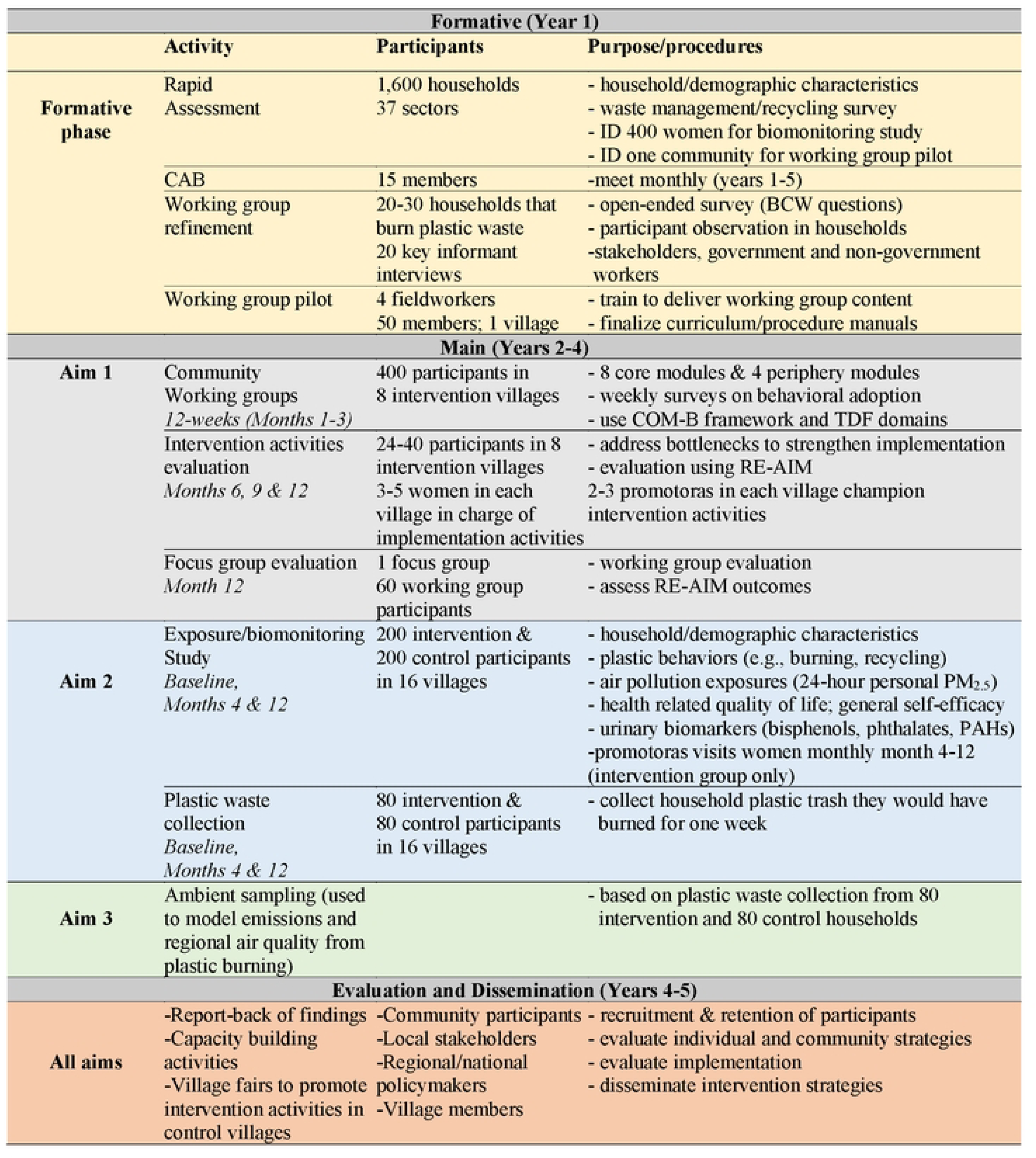
Ecolectivos summary of study activities and timeline

### Surveys

Data for the outcome assessments will be obtained directly from participants through questionnaires, interviews, or observations. During scheduled home visits over a period of two days, fieldworkers will verbally administer questionnaires in Spanish about: (a) sociodemographic characteristics; (b) location and frequency of burning waste, including plastic; (c) types and amount of waste that is burned or recycled; (d) sources of household smoke, including stove type and tobacco exposures; and (e) food/products containing Polycyclic Aromatic Hydrocarbons (PAHs), Volatile Organic Compounds (VOCs), Bisphenol A (BPA) and phthalates that will be measured in the biomonitoring study. Additionally, we will administer five instruments to measure changes in women’s health and well-being, specifically Health-related Quality of Life instrument [37], Household Decision Making [38], New General Self-Efficacy Scale [39], Short Social Capital Assessment Tool [40], and the Community Mobilization Scale [41] **(Table 3)**.

**Table 3:** Study instruments to measure women’s health and well-being.

|  |  |
| --- | --- |
| CDC Health-related Quality of Life (HRQoL) instrument [37] | 4 items: 1) would you say that in general your health is excellent, very good, good, fair, or poor?; 2) for how many days during the past 30 days was your physical health not good?; 3) for how many days during the past 30 days was your mental health not good?; and 4) during the past 30 days, how many days did poor physical or mental health keep you from doing your usual activities? |
| Self-Efficacy [39] | Self-Efficacy will be measured using the validated New General Self-Efficacy Scale. It utilizes an 8-item scale that is measured by a 5-point Likert scale, ranging from 1=strongly disagree; 3=neither agree nor disagree; 5=strongly agree. |
| Household Decision Making [38] | Household decision making will be measured by household decision making scale. This scale utilizes 7 questions, and the responses are coded on 4-point scale: 1=Husband. 2=Joint decision, 3=respondent decision or others. |
| Community Social Capital [40] | Community social capital will be measured using a validated measure Short Social Capital Assessment Tool (SASCAT). |
| Community Mobilization [41] | Community mobilization will be assessed using two subscales (collective action and critical consciousness) from the validated community mobilization measure. The responses are noted on 3-point Likert scale agree, disagree, somewhat agree. |

### Estimates of plastic trash burning

From each village, we will randomly select 5 participating households and collect all nonorganic waste, including plastic. We will instruct them to collect all household trash produced over a 1-week period, at baseline, 4 months, and 12 months, in bags we provide. After fieldworkers sort and weigh the plastic waste at the field office, trash will transported to the municipal dump and/or the recycling center in Jalapa City. These estimates will be used to capture changes over time in plastic waste that would have burned and used for modeling plastic emissions estimates in Aim 3.

### Air pollution field sampling

#### Personal sampling

We will use air pollution sampling techniques employed in previous studies [42–44]. We will assess exposures to personal airborne PM_2.5_ and BC in all 400 women. In a subset of 60 women, we will measure 1,3,5-TPB and a series of 25 PAHs on quartz filters twice (60 at baseline and 60 at 4 months; 30 each from intervention and control). We will use a personal monitor placed in apron pockets that we provide to participants. We will instruct women to wear the apron throughout the day and to place it near their bed at night. On day 2, we will retrieve the equipment and administer a survey about daily activities/compliance. We will use the Ultrasonic Personal Air Sampler monitor (UPAS V2 Plus, Access Sensor Technologies, Fort Collins, CO) which provides both real-time and filter-based PM_2.5_. The UPAS is lightweight, quiet, durable, and has a battery that lasts over 24 hours. The device uses a cyclone for PM_2.5_ size separation, and a small pump that maintains a constant flow rate, measured internally using pressure sensors. The device is configured over Bluetooth using an iPhone app [45].

#### Kitchen and outdoor fire air pollution sampling

In 80 kitchens we will measure PM_2.5_ using the UPAS at the same time as personal exposure assessment and urine collection. The kitchen UPAS monitor will be affixed to a kitchen wall 150 cm from the floor and 100 cm from the center of the fire. For open fire monitoring, 2 tripods with UPAS monitors will be set up (1 with quartz and 1 with PTFE filters), 1 meter from the outdoor fire. The fire will be monitored at a high flow rate (3L/min) for 60 minutes. We will note behaviors related to starting, stoking and if a woman who is wearing personal monitors is standing by the outdoor fire.

We will analyze PM_2.5_, BC on PTFE 37 mm filters and 1,3,5-TPB and PAHs on quartz 37 mm filters. Prior to deployment in the field, PTFE filters will be pre- and post-weighed in the air pollution laboratory at the Universidad del Valle de Guatemala using a microbalance (Sartorius Cubis MSA 6.6S-000-DF-00) to calculate the gravimetric PM_2.5_ concentrations. A SootScan™ (Model OT21 Optical Transmissometer) will be used to analyze BC. The quartz filters will be pre-baked at 550 °C at the Stone Research Group Laboratory at the University of Iowa. Each week, there will be one lab blank sample that will be included for both quartz and PTFE filters and in laboratory settings.

#### Ambient monitoring

With the permission of the Guatemalan Ministry of Health officials, we will set up local PM_2.5_ ambient monitors (MODULAIR-PM, produced by QuantAQ, Inc, Somerville, Massachusetts) on the rooftops of four health centers in participating villages, selected for their distance from main roads and stable electricity supply. A fifth monitor is located on the field station in Jalapa city. The MODULAIR-PM measures size-resolved particle number concentrations and particulate matter (PM) at different diameters, as well as temperature and relative humidity. Data is transmitted via cellular phone data services and can be viewed globally via the QuantAQ Cloud platform.

### Air pollution specimen analysis

#### Particulate matter

(PM_2.5_): Before and after sample collection, PTFE filters will be conditioned and weighed at the Universidad del Valle de Guatemala (UVG) filter weighing lab. PM_2.5_ mass will be calculated as the difference between pre- and post-sampling filter weights, each determined in duplicate.

PM_2.5_ masses will then be converted to mass concentrations by dividing by the sampled air volume. Mean change in field blank filter masses (5% of samples will have field blanks) will be subtracted. We will measure precision by performing duplicates in 5% of samples.

#### Black carbon

At the UVG lab, we will analyze 1,200 filters to determine the optical attenuation and black and brown carbon mass. The analysis is non-contact, non-contaminating and non-destructive. Therefore, filters can be subsequently analyzed for PAHs/VOCs and metals. The analysis method requires no support gases or consumables.

#### PAHs

A subset of 120 37-mm quartz filters will be analyzed at University of Iowa to assess the composition and mass of TPB and 25 PAHs, as done in previous studies [48]. The remaining quartz filters will be stored for future analysis when funding is available. Extracted samples will be analyzed by gas chromatography (GC; Agilent Technologies 7890A) coupled to mass spectrometry (MS; Agilent Technologies 5975). All measurements will be blank-corrected.

### Urinary Biomarkers

We will use urine measurement techniques that were successful in previous studies [43,44,49]. On day 1, when air pollution exposure sampling begins, female fieldworkers will instruct women on clean-catch procedures and provide a sterile urine collection cup and vaccine cooler with ice packs. Women will be instructed to collect a first-morning void sample on day 2. At the home visit on day 2, fieldworkers will administer surveys, including collection time. Labelled specimens will be secured in a vaccine cooler and transported in project vehicles to the field laboratory. Urine will be refrigerated within four hours of collection.

In the field laboratory, 21 ml of urine will be transferred to four cryovials (two 4 ml tubes with 3 ml urine; and two 10 ml tubes with 7.5 ml urine) using QR code labels that link to the participant ID. Samples will be stored at -20°C in the field laboratory and transported to the Universidad del Valle de Guatemala laboratory within 3 months. They will be stored in -80°C freezers at the UVG laboratory until shipped with freezer packs to the Barr Lab at Emory where they will be stored at -80°C until urinalysis is performed. We will not include information about the study arm so that blinded data will be analyzed. If samples are accessed by investigators other than those named in this protocol, data will be requested by the investigators using a data request form and reviewed by the MPIs before approving any requests.

### Urine biomarker specimen analysis

Target analytes are outlined in **Figure 3**. Eight PAH and six VOC analytes will be targeted. 1.0 ml (PAHs) and 0.5 ml (VOCs) of urine will be aliquoted and spiked with isotopically labeled internal standards for automatic recovery correction and normalization of mass spectral data. Nine phthalate and two bisphenol analytes will be targeted. A 1.0 ml aliquot of urine spiked with isotopically labeled analogues of target phthalates and phenols will be subjected to enzyme hydrolysis to liberate glucuronide-bound conjugates. The concentrated extracts will be analyzed using GC-MS/MS. Two quality control materials (one high and one low) and one blank sample will be analyzed concurrently with each set of 28 unknown samples, and NIST reference standards will be used for quality assurance measures.

**Figure 3.**
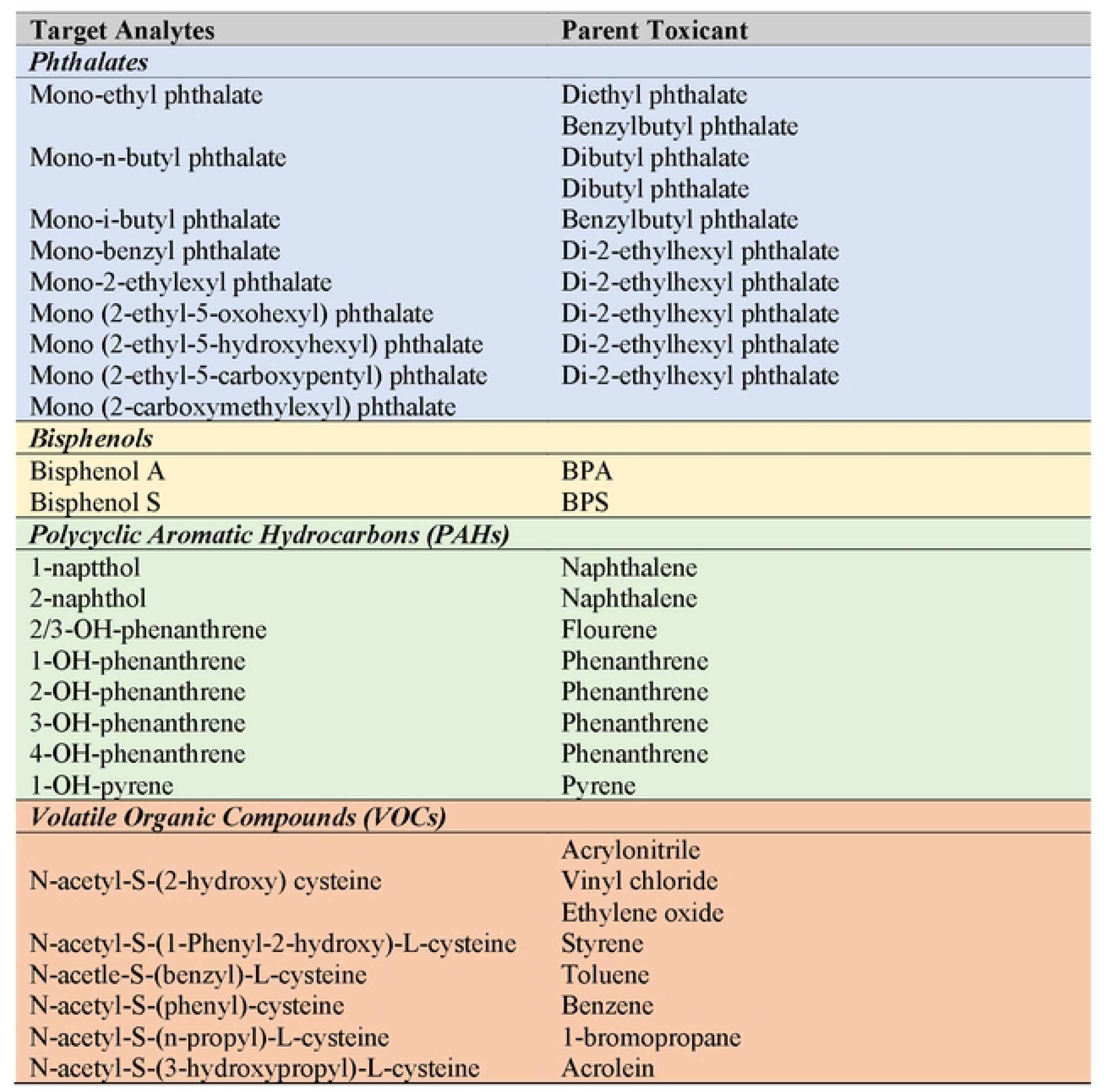
Target analytes and parent toxicants

### Focus groups to assess intervention activities

In each intervention village, we will organize meetings in each community to assess the sustainability of interventions that were chosen over the 9-month follow-up period. These meetings will occur at 3, 6, and 9 months after the community working groups have ended. The purpose of these meetings is to troubleshoot problems that are impeding the success of the chosen interventions and offer supports from project staff as needed. We will also conduct focus groups to evaluate the working groups at 13 months, inviting 6-8 women (n=60-80) who did not participate in the activities. The purpose of these focus groups is to evaluate the reach of the chosen interventions among those who did not participate in intervention activities.

### Data Quality

The Dual-PIs and co-investigators at UVG will oversee personnel training, including standardization and data entry/management. The quality of air pollution and urinary biomarker monitoring will be assured during lab analysis, specimen handling and calibration at the field headquarters, and protection of specimens in transport. Standard operating procedures (SOPs) will be developed in English and Spanish. All survey forms will be pilot tested, translated into Spanish, and back translated into English to confirm consistency with original questions.

### Retention

#### Control villages

Each participating household will receive 25 sapling trees to plant on their land. Community leaders of the eight control villages will receive 300 sapling trees for local reforestation. Community leaders will choose areas to be reforested and will organize the activities with study staff.

#### Intervention villages

We will seek permission form local village leaders to support retention of women in the working groups. Study staff will seek solutions to encourage regular participation in working groups (e.g., securing a meeting room, providing transportation and childcare). We will use community-engaged research to develop intervention activities that are chosen by the working group participants. Promotors will visit participating women to assess their engagement in intervention activities.

## Data Management

Data management will be overseen in Guatemala by the project manager and the data manager. Reports will provide summaries such as the number of visits completed, exceptions to the protocol (missed visits, missing forms), and dates of expected visits. Summaries on completeness will be provided. Survey forms, data dictionaries and databases will be created in REDCap (Research Electronic Data Capture), Participant identifiers will be stripped of protected health information and will be assigned in a way to improve tracking and monitoring of study activities. Staff will use pre-printed QR-coded ID labels to avoid transcription errors when labeling study samples.

## Data Analysis

Baseline assessment and working group refinement surveys will be analyzed using descriptive statistics. Key informant interviews will be audio-recorded and transcribed. Transcripts, notes from participant observations, and ethnographic field notes will be coded and analyzed. We will use thematic analysis, which is flexible and a theoretical and can be applied across a range of qualitative methodologies. Categories (i.e. themes or variables) and their properties (sub-categories) will be tested deductively [51] based on the constructs of the BCW. New themes will be identified and coded using an inductive thematic approach.

Mixed methodologies offer causal explanations grounded in different kinds of empirical data [52]. Informant-driven qualitative findings, such as focus group responses post-intervention, will be: 1) assessed against demographic characteristics of working group participants; 2) linked to the reach, effectiveness, adoption, and sustainability of intervention strategies (Aim 1); and 3) compared to quantitative findings from air pollution exposures and biomarkers (Aim 2). Thus, mixing methods may contextualize both the depth (qualitative) and breadth (quantitative) of patterns of plastic burning and changes related to the intervention strategies [53].

## Aim 1: Evaluation of intervention activities using implementation research approaches

Our evaluation of intervention implementation involves components of the COM-B model/TDF and the RE-AIM framework. Measures related to the COM-B model (described previously) that are included in the study data collection are outlined in **Table 2**. The evaluation of the intervention includes three main approaches: (1) we will measure individual-level changes in plastic waste use and burning behaviors among all working group participants over the age of 15 years of age using survey methodologies and field notes (intervention arm only); (2) we will evaluate the community-level changes in these same plastic-use and burning-related measures among the 400 participants involved in the biomonitoring study, also with survey methodology (both intervention and control arm); and (3) we will measure changes in urinary biomarkers and personal exposures to air pollution in 400 among women in the biomontioring study (both intervention and control arm). Together these approaches will inform us about community-level impact among women who attend and don’t attend the working groups.

We will also use the RE-AIM framework [65,66], primarily focusing on overall reach as well as on effectiveness, as measured by changes in individual-level household behaviors (women who participate in working groups in intervention villages). Specifically, *Reach* of the intervention will be assessed by participants’ level of engagement in working groups (attendance frequency, duration and completion). *Adoption* will be measured by the number and types of behavior changes based on working group strategies. This will include the number of participants in community working groups who report reducing, reusing, or recycling plastic and reduced plastic burning, as reported by survey data. Additionally, we will measure village-level reach measured by the level of involvement in village level activities that arise from working group survey data [67]. *Effectiveness* will be measured by the level of behaviors (high/low levels of reducing, reusing or recycling plastic) reported by participants and its effect on perceived health and well-being outcomes, including collective efficacy, general self-efficacy, health-related quality of life, and reductions of urinary biomarkers of exposure. Using monthly data collected at the household level by promoters, we will assess *Maintenance* by the number and proportion of intervention participants (25 biomonitoring women and working group attendees) who remained engaged in one of more plastic waste reducing behavior (e.g., participation in a community clean-up program) to reduce consumption, increase recycling, and who report no longer burn plastic. We will assess which program components are essential for success [68]. Results for Aim 2 will be analyzed and disseminated by May 2027.

Aim 2: Longitudinal effect of behavioral intervention on air pollution and urinary biomarkers

### Urinary biomarkers and Air Pollution

Urinary biomarker and air pollution concentrations are typically right-skewed and will be described as the geometric mean (95% confidence interval). We will estimate variance components to assess intraclass correlations among repeated measures within subject and village.

### Analysis Methods

We will log transform our data and use linear mixed effects models. Multi-level mixed effects models (MLM) with random (individual- and village-level) and fixed (treatment group, study period, and group- by-period interaction) effects will be employed to test for trajectories of change for intervention group versus control group focusing on group-by-time interaction effect. If there are significant differences between the 2 groups at baseline (e.g. difference in ambient air pollution), these will be controlled by design and estimated by the main effect for study group. MLM longitudinal models evaluate the changes across the three time points, and interaction of group and each separate follow-up time point will allow planned post hoc estimates of both the immediate intervention effects (4 months) and the longer term sustained effects (12 months). To adjust for potential confounding, as well as to increase efficiency by reducing residual variance in urinary biomarker concentrations, we will adjust for indicators of dietary intake (ingestion route) and cosmetic product use (dermal route) plastic contaminants. We will use the Stepdown-minP procedure to calculate p-values that take into account multiple comparisons and the correlation expected among multiple measures of air and urine concentrations [55]. All model assumptions will be tested with standard diagnostic tests and influence statistics used to test the distributions of the residuals. MLM utilize all available data for all participants at each time point.

### Heterogeneity of intervention effects

In addition to overall effect, we will explore heterogeneity in effects between communities using a meta-analysis approach. This will be done by estimating the intervention effect within each of the matched-pair communities, aggregating the effect sizes across matched pairs to derive an overall intervention effect, and testing for heterogeneity in the effect of the working group interventions on exposure to products of plastic combustion across matched pairs of communities [49]. This examination of variation in the intervention effect across matched-pairs has the potential to answer important questions about why the working groups may be more successful in one set of matched pairs compared to other matched pairs. For example, the heterogeneity may be partly explained by fidelity to the working group protocols, the level of community participation, or the types of interventions that are implemented. Moreover, as a secondary analysis we will explore the impact of the interventions not only on the mean of the exposure distribution but also on its shape. This will be performed using quantile regression to compare distributions between and within groups [54]. This will allow us to compare the effects of the interventions at various locations of the exposure distribution.

### Missing Data

From our previous experience conducting longitudinal studies in rural Guatemala, we expect about 85% completeness accounting for loss to follow-up and intermittent missingness due to logistic, communication and technological failures [56]. We will use the doubly robust estimation method to address bias due to missing data [57]. First, we will explore models to predict missing exposures, including predictors such as baseline characteristics, previous exposure measures, and reported and observed behaviors related to exposure sources. If this model has predictive validity, we will use it in a multiple imputation sensitivity analysis. Our second approach will be to build a model that explains missingness by modelling the probability of missingness conditional on treatment assignment and baseline covariates, as in inverse probability weighting. Finally, if both models are found to be predictive, we will apply the doubly robust estimation for targeted inference, which relies on parameters from either of these models being estimated consistently [58]. R software will be used [59]. Results for Aim 2 will be analyzed and disseminated by May 2027.

## Aim 3: Model regional air pollution scenarios from plastic waste burning

We will collect all trash that is produced by 10 participating families in each village and estimate masses of plastic trash to quantify emissions of 62 chemical species, as described in Bardales et al. [32]. The emissions estimates for each of the species will have uncertainties associated with them by conducting 1 million Monte Carlo samplings using the mean and standard deviation of mass estimates and emission factors (EFs). Using the most recent 2018 Guatemalan census data and the amount of garbage produced [60], as well as the percentage of plastic waste from the recent World Bank database [61], we will create emissions estimates, again using the 1 million Monte Carlo samplings. Due to data availability for Guatemala, in addition to Jalapa, we have city-level emissions estimates for Guatemala City, Antigua, and Jutiapa. For other parts of Guatemala, we will use country-level estimates. These emissions estimates will then be spatially distributed across Guatemala, following the garbage burning emissions estimates distribution in our pilot study [32]. This activity will occur three times at baseline (the start of the study), 4-6 months and 12-13 months from the baseline visit.

For forecasting future emissions from plastic burning, we will create several emissions scenarios for the year 2030. Based on differences in plastic waste collected in households over time, we will estimate the mitigation potential for different chemical species of interest from reduced plastic waste in Jalapa, Guatemala, following the similar methodology used in Saikawa et al. (2020) [62]. We will then use the “online” Weather Research and Forecasting (WRF) model coupled with Chemistry (WRF-Chem) [63,64] to simulate the regional air quality over Guatemala and Central America to assess the impacts of different plastic emissions on air quality at the local and regional levels. The model domain will cover most of the Central America region with two nested domains: one for Guatemala and the innermost domain for Jalapa. For each scenario, we will use WRF-Chem to model air quality for several months of the year to understand seasonality. For each simulation, the model will be spun up for 14 days to allow the model to ventilate the regional domain and this period will not be included in the analysis. For our baseline simulation, we will evaluate our WRF-Chem model results by comparing simulated pollutant concentrations with observational data available in Guatemala. Results for Aim 3 will be analyzed and disseminated by May 2027.

## Methods: Monitoring

The potential risks from data collection procedures are minimal. Most data are obtained by interviewer-administered surveys, observations, or non-invasive procedures and pose no risk of physical harm, and therefore we do not have an independent data monitoring committee. Reporting of adverse events will follow requirements established by the Institutional Review Boards at Emory and UVG. Data will be reviewed and reported to the Dual-PIs at Emory and UVG by the field project manager at the research site.

## Ethics and dissemination

### Research ethics approvals

This study has approval from the Emory University Institutional Review Board (#00002412) and the Research Ethics Committee of the Center for Health Studies of the Universidad del Valle de Guatemala (approval # 245-05-2021). Reliance agreements are in place with the University of California San Francisco and the University of Georgia to rely on Emory’s IRB approval. All study documents including approval letters and protocol amendments will be communicated to study investigators made available via a secure web-based document-sharing platform (e.g., OneDrive).

### Informed consent

Written informed consent or assent for all procedures, including ancillary studies will be obtained by a trained local fieldworkers in Spanish. Steps will be taken to ensure participants’ comprehension at an elementary-school level. Written assent will be provided for participants between 15 and <18 years of age, and the designated legal guardian will provide consent for their child to participate in the study. The study will employ standard methods for protecting the confidentiality and privacy of participants.

### Access to data

Access to projects will be assigned by the Dual-PIs. Access to data files is strictly monitored by the project manager. All requests for data use will be approved by the Dual-PIs. Public access to a de-identified dataset and statistical code will be published and freely accessible in a data repository using a digital object identifier (DOI) for other researchers’ use.

### ***A***ncillary and post-trial care

The potential risks from data collection procedures used in this study are minimal. Most data are obtained by interviewer-administered surveys, observation, or non-invasive procedures conducted during the biomarker and air pollution monitoring components of the study and pose no risk of physical harm to the participant or other household members. There are no provisions for proposed ancillary of post-trial care for this study beyond usual care in local health facilities.

### Dissemination Policy

Results will be shared with participants and participating communities at dissemination meetings conducted in the control communities in Year 5. We will discuss successful practices intervention communities developed to reduce plastic trash burning as a result of the workshop activities (Aim 1). We will invite local policymakers to attend meetings to learn about the results. We will evaluate changes in expousres to air pollutants and urinary biomarkers in women measured at 4 and 12 months after baseline (Aim 2). Aggregated results will be presented at dissemination meetings in Year 5. We will also present air quality modeling results (Aim 3) at the dissemination meetings. We will present findings in such a way that they are understandable to lay audiences. Results will be written by co-investigators, with authorship based on the International Committee of Medical Journal Editors (ICMJE) guidelines [69] and presented in open access, peer-reviewed scientific journals and at scientific conferences and will involve collaborators as appropriate.

## Discussion

Plastic waste disposal, including incineration, are major problems in waste management [70]. To mitigate this problem, local, national and international policies need strengthening. While there are global efforts to introduce clean cookstoves, these programs do not address plastic that is commonly burned in household fires. Aside from our own research to estimate plastic waste burning [62] and our pilot study for this proposal [71], few intervention studies have examined community efforts to limit household burning of plastic waste. Furthermore, there are no global emissions estimates from plastic waste burning in developing countries where a majority of households burn waste as the primary means of disposal [60].

This is the first study to use a behavioral framework to systematically refine and implement an intervention to address plastic waste burning, as well as conduct detailed exposure assessment. We will use a village-level cluster randomized design to evaluate whether community/individual behavioral interventions reduce the quantity of plastic waste combustion. We will refine, implement and evaluate the reach, effectiveness, adoption and maintenance of intervention strategies from village-level working groups using the RE-AIM framework. Using a randomized design will allow us to evaluate the causal effects of these interventions on women’s personal exposures and urinary concentrations of contaminants produced by plastic combustion. This study will provide estimates on how much plastic waste is burned in homes in Jalapa, Guatemala, and how this is manifested in air pollution exposures and urine concentrations of chemicals found in women exposed to burning plastic. We will create several emissions scenarios, based on our estimates and census data, to assess the impact of the intervention to reduce plastic waste burning on local and regional air quality.

The long-term goal of the Ecolectivos study is to build sustainable local and national strategies based on findings from community working groups to reduce air pollution from plastic trash burning. This has implications for other low-resource communities where plastic waste is burned in informal trash fires. Our findings may inform implementation policies in low-income countries to improve community responses to air pollution from these sources. By modeling contributions to air pollution made by plastic burning in one region, we provide an approach for future intervention programs that combine behavioral intervention evaluation with exposure assessment for policy-relevant solutions.

## Supporting Information

**S1:** SPIRIT 2013 checklist: Recommended items to address in a clinical trial protocol and related documents.

## Data Availability

No datasets were generated or analysed during the current study. All relevant data from this study will be made available upon study completion.

